# Factors associated with the transition from health to disease for type 2 diabetes, cardiovascular disease and kidney dysfunction: A NAKO-IMMEDIATE analysis

**DOI:** 10.64898/2026.10.01.26364547

**Authors:** Solange Gonzalez Chiappe, Katharina Nimptsch, Ilais Moreno Velásquez, Martina Bouka, Heiko Becher, Marcus Dörr, Michael Leitzmann, Henry Völzke, Sabine Schipf, Stefan N. Willich, Thomas Keil, Peggy Sekula, Karin Halina Greiser, Justine Tanoey, Clara Frick, Annette Peters, Julia Six-Merker, Gertraud Maskarinec, Tobias Pischon, the IMMEDIATE project

## Abstract

**Background:** Cardio-kidney-metabolic (CKM) diseases share risk factors, but their shared and distinct risk factors for disease progression remain unclear. We examined whether classical risk factors predict baseline CKM indicators and their longitudinal changes in a population-based German National Cohort (NAKO) sample.

**Methods:** In 2,650 NAKO-IMMEDIATE participants selected for deep-phenotyping, baseline values and 5-year changes in — glycated hemoglobin (HbA1c), systolic blood pressure (SBP), pulse wave velocity (PWV), and estimated glomerular filtration rate (eGFR) were linearly regressed on age, sex, education, alcohol and tobacco consumption, body mass index (BMI), and residuals of BMI-adjusted waist circumference (WC) with mutual adjustment.

**Results:** Baseline values for HbA1c were 34.6±5.8 mmol/mol, for PWV 8.3±1.5 m/s, for SBP 128.0±15.7 mmHg and for eGFR 96.0±14.6 ml/min/1.73 m². Cross-sectionally, all risk factors were associated with one or more of the CKM indicators, with age and BMI being associated with all four indicators. Over 5 years, HbA1c (1.2±4.2 mmol/mol), PWV (0.5±1.2 m/s) and SBP (0.6± 14.2 mmHg) increased, whereas eGFR (−2.3±9.5 ml/min/1.73 m²) decreased. Age (0.03 m/s per 5 year, 95% CI 0.01, 0.06 m/s) was associated with higher and BMI (−0.02 m/s per 1kg/m2, −0.03, −0.01) with lower PWV changes. No other risk factors were associated with CKM indicator changes.

**Conclusion:** Classical risk factors were more frequently related to baseline CKM indicators than to their 5-year changes. Within longitudinal analyses, only age and BMI were related to PWV 5-year changes.

**Clinical Perspective:**

- It is well established that classical risk factors are associated with cardio-kidney and metabolic (CKM) diseases whereas their associations with longitudinal changes in CKM indicators are less well understood.
- This study examines to what extent age, sex, education, alcohol and tobacco consumption, BMI and waist circumference are associated with glycated hemoglobin, systolic blood pressure, pulse wave velocity and estimated glomerular filtration rate at baseline and their 5-year changes.
- Only age and BMI were associated with changes in PWV, while other longitudinal associations were non-significant, suggesting that classical CKM risk factors have greater utility for assessing cross-sectional CKM function than for predicting prospective changes in CKM function in the general population.

## Introduction

Cardio-kidney and metabolic (CKM) diseases remain the primary cause of disease burden worldwide^1^. They constitute a group of intertwined disorders that include cardiovascular disease, type 2 diabetes, chronic kidney disease and other conditions. Although typically diagnosed during the second half of life, they are often preceded by a progressive organ function decline that might have begun years before^2^. Tracking this organ function decline at early stages and studying its determinants may convey useful information on CKM disease risk onset and on potential preventive measures. Hemoglobin A1c (HbA1c), pulse-wave velocity (PWV), blood pressure (BP) and estimated glomerular filtration rate (eGFR) are common indicators of type 2 diabetes cardiovascular disease and kidney function, respectively, and can be used to track organ functions^3–6^.

Numerous cross-sectional studies have reported associations of classical risk factors, namely age, sex, education, alcohol and tobacco consumption, body mass index (BMI), and waist circumference (WC) with these CKM indicators. For instance, higher age, male sex, low educational level, higher BMI or higher WC have been usually associated with higher HbA1c^3, 7, 8^, higher PWV^9, 10^, and higher BP^11, 12^. Higher age, higher BMI and higher WC have also been consistently associated with lower eGFR^13–15^. Finally, alcohol^16–18^ and tobacco consumption^7, 19^ have been related with higher BP or higher HbA1c.

However, there is less evidence regarding the extent to which classical risk factors also predict changes in HbA1c, PWV, BP and eGFR; beyond the average CKM function decline over time. For example, while it is well established that people with higher BMI have higher HbA1c concentrations^8^, it is less clear whether higher BMI is also associated with a stronger prospective increase in HbA1c^3, 7, 20^. Males have usually higher mean SBP^5^ or higher mean PWV^4^ than females; however, a stronger prospective change of those cardiovascular indicators over time has been described in females^21^. In addition, while there is evidence that GFR varies with age, sex or BMI^6^ inconsistent associations were reported for age and sex with GFR decline over time^22^. Information on risk factors of changes in CKM function is important to gain a deeper understanding of the transition from health to disease but also for the planning of interventions on modifiable risk factors aiming at disease risk reduction.

We therefore used data from the IMMEDIATE (Imminent Disease Prediction and Prevention at the Environment Host Interface) EU project within the German National Cohort [NAKO]^23^ to examine to what extent classical phenotypical, demographic and lifestyle risk factors predict CKM function indicators at baseline and their longitudinal changes in a NAKO sample.

## Methods

### Study Population

The NAKO (DRKS00037328) is an ongoing and prospective large cohort, randomly sampled from the general population, designed to investigate risk factors, mechanistic pathways, markers of early detection, and risk prediction for a broad range of diseases^24^. The recruitment process and data collection have been described previously^25^. Briefly, from 2014 to 2019, NAKO enrolled 205,415 adult participants 19 to 74 years of age at 18 study centers across Germany, among whom 10,085 were enrolled at the NAKO Berlin-North center. Participants were randomly selected from population registries. The design intended to recruit for both women and men 10.0% of participants in each 10-year group between 20 and 39 years of age, and 26.7% in each 10-year group between 40 and 69 years of age. From 2019 to 2024 (4 to 5 years after baseline), all NAKO participants were re-invited to a 5-year follow-up examination and 137,922 participated (among them 7,425 in the NAKO Berlin-North center). Both baseline and 5-year examination included personal computer-assisted standardized interviews, self-administered questionnaires, in-depth biomedical examinations including collection of a wide range of biosamples and laboratory measurements. The NAKO has been approved by the relevant ethics committees, and written informed consent was obtained from all participants^26^.

For the current analysis, a random sub-sample of 2,650 participants from the NAKO Berlin-North center (aged 20-71) was selected within the context of the IMMEDIATE project (Figure 1). This project, created for deep omics phenotyping, prioritized participants with available stool samples at baseline (stool-derived multi-omics profiles) and/or participants who had a brain magnetic resonance imaging at both baseline and 5-year examination (for neuroimaging markers of brain health).

**Figure 1.**
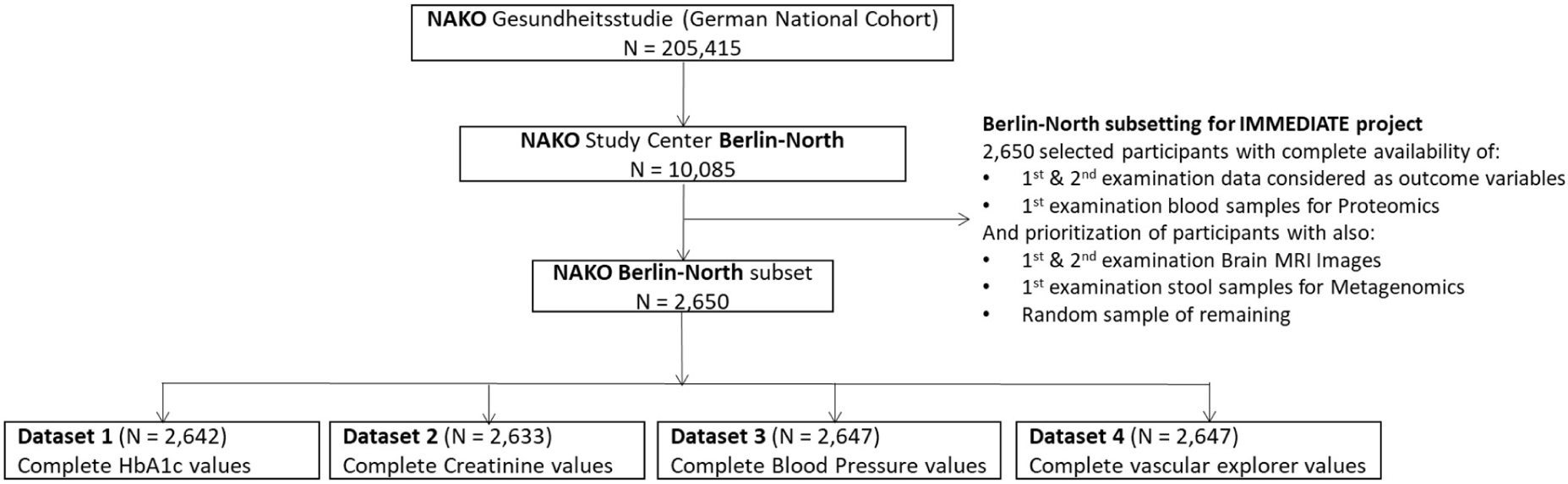
Flow-chart from the NAKO sample at a national level to a Berlin-North for NAKO/IMMEDIATE subset and to four datasets with complete outcome data

### Classical risk factor assessment

Age, sex and educational attainment were obtained by face-to-face interview. Educational attainment was assessed via the ISCED 97 (International Standard Classification of Education 1997) levels: “Low”, “Medium”, “High” educational attainments and “Enrolled in education”. Alcohol and tobacco consumption were obtained by self-administered touchscreen questionnaire. Alcohol consumption was categorized in: “Never”, “Former”, “Current alcohol consumption between 0 and 5g/day”, “Current alcohol consumption between 5 and 15 g/day” and “Current alcohol consumption of more than 15 g/day”. Tobacco smoking status was categorized in: “Never”, “Former” and “Current” smokers. BMI was computed with participant’s measures of height (SECA Stadiometer 274) and weight (SECA Medical Body Composition Analyzer mBCA 515, SECA, Hamburg, Germany) as kilograms per meter squared (kg/m²). The SECA Circumference measuring tape 201 was used to measure the WC (at an accuracy of 0.1 cm).

### Cardio-kidney and metabolic function indicator assessment

The HbA1c was measured in mmol/mol in EDTA plasma by High-Performance Liquid Chromatography with D-100 and Variant TM II TURBO Hemoglobin Testing Systems (Bio-Rad Laboratories, Hercules, USA). The SBP was measured in mmHg twice in the sitting position after 5 min of rest and recorded at 2-min intervals using an Omron Hem-705IT device (the second measurement was used for analyses as proposed by Schikowski et al.^27^). Brachial-ankle PWV was measured using the Vascular Explorer® medical device (Enverdis GmbH, Germany). It was assessed by placing blood pressure cuffs around the left upper arm and the left ankle and inflating them to diastolic BP for 15 s. During this time the pulse wave was recorded by oscillometer, and brachial-ankle pulse transit time (baPTT) was measured by device-specific algorithms detecting the real time foot-to-foot difference between brachial and ankle pulse wave. The distance between the center of the blood pressure cuff on the arm and the suprasternal notch (L1) and the distance between the suprasternal notch and the middle of the blood pressure cuff on the ankle (L3) were measured using large plastic dividers. PWV was estimated by dividing (L3-L1) by baPTT^28^. We selected brachial-ankle PWV over carotid-femoral PWV due to its higher reliability in a repeated measures study design and with measured distances over calculated distances due to its higher accuracy^28^.

Estimated GFR (eGFR) was assessed with the CKD-EPI 2009 formula that combines information on serum creatinine, age and sex of the participant^29^. Serum creatinine was measured in µmol/l by photometry (Jaffé) with AU5800 and AU680 (Beckman Coulter, Brea, USA) in a local laboratory. Within the selected sub-sample, baseline and follow-up CKM indicators were assessed using the same corresponding procedures and medical devices.

### Statistical analyses

Normally distributed data was summarized by their mean ± standard deviation (±SD), whereas non-normally distributed data was summarized by their median and 25-75% interquartile range (IQR). Counts were expressed as frequencies and percentages (%). Age was centered at 50 years. We used separate multivariable linear regression models with robust variance estimation to study the cross-sectional associations between the baseline classical risk factors and the baseline CKM indicators and the longitudinal associations between the baseline classical risk factors and the CKM indicator 5-year changes. Each risk factor was considered as an exposure in the multivariable models, i.e. all risk factors were mutually adjusted. The WC residuals were obtained by regressing WC on BMI, and used as predictors in the linear regressions to avoid multicollinearity. Five-year changes in CKM indicators were calculated for each participant as the difference between the follow-up examination and the baseline values divided by the time interval between these examinations, multiplied by 5 years. For illustration, we also categorized changes in CKM indicators relative to baseline values: >10% (increase from baseline value), −10% ≤ change ≤ 10% (no substantial change), and <10% (decrease from baseline value). These thresholds were set as approximations of established reference change values of repeated measurements that differentiate true clinical changes from random fluctuation at an individual level^30–32^. Longitudinal analyses (*change score models*) were not adjusted for baseline outcome values due to the risk of inducing spurious associations between the exposure and the studied outcome. This is particularly relevant for extreme baseline values which are often associated to unmeasured or unknown confounders and/or to measurement errors; controlling for them in observational non-randomized studies can lead to biased results^33, 34^. For each multivariable linear regression, datasets with complete outcome data were used (Figure 1). Missing values for independent variables were handled using multiple imputation with fully conditional specification (FCS) in PROC MI (SAS), generating 10 imputed datasets, with estimates combined using PROC MIANALYZE according to Rubin’s rules.

As sensitivity analyses, linear regressions were further adjusted for prevalent self-reported diabetes and hypertension medication and also for total physical activity per day in two different models. In the NAKO cohort, a diabetes or hypertension diagnosis, as well as medication use, are informed by participants during the face-to-face interview where they bring medication packaging. In order to maximize diagnosis accuracy, only medication use was incorporated as exposure variable in the linear models. Notably, as medication uses were considered intermediate variables in the association between classical risk factors and CKM indicators, they were not confounding the total effect association and therefore were included in sensitivity analyses uniquely. Total physical activity per day was assessed by personal computer-assisted standardized interviews along the General Physical Activity Questionnaire (GPAQ). It was assessed in minutes per day and was winsorized for values >960 min/day. In sensitivity analyses, linear regression models were stratified by sex, and sex-by-exposure interaction terms were tested in the overall sample using Type III tests of fixed effects in PROC MIXED (SAS Enterprise Guide 8.4) to assess whether exposure – outcome associations differed by sex.

Analyses were conducted using SAS Enterprise Guide, version 8.4 (8.4.1.15; SAS Institute Inc., Cary, NC, USA). A false discovery rate (FDR) procedure was applied for the cross-sectional and longitudinal main multivariable linear regression results. Estimates were considered to be statistically significant when P values adjusted for False Discovery Rate obtained with the Benjamini-Hochberg procedure were < 0.05. For categorical exposure variables, statistical significance was assessed using P values from Type III tests of fixed effects in PROC MIXED, based on the fully adjusted models, providing an overall test of the association between each categorical exposure and the outcome. In sensitivity analyses, statistical significance of categorical exposure variables was assessed for each dummy variables obtained from linear regressions.

## Results

### Baseline characteristics

Among the 2,650 participants, median age was 50.0 years (IQR 43.0,59.0) and females represented 48.6% of the sample. Age distribution was bi-modal by design (participants older than 40 years were over-represented). The majority of participants were of European ancestry (99.2%), had medium and high education levels (97.3%), were current alcohol consumers (94.5%) and never or former smokers (83.2%). Median BMI was 25.5 (IQR 23.0,28.5) kg/m^2^, (24.6; 22.1,27.8; kg/m^2^ in females, 26.3; 24.2,29.0; kg/m^2^ in males). Median WC was 88.8 (IQR 80.1,98.3) cm, (83.7; 75.2,90.3; cm in females, 94.8; 87.1,102.4; cm in males) (Table S1). Hypertension, diabetes mellitus and medication for hypertension and diabetes were self-reported in 28.0%, 5.2%, 23.8% and 3.1% of participants, respectively. Missing data was ≤1.1% for alcohol, tobacco consumption, WC and hypertension and diabetes medication variables; it was of 7.6% for educational attainment (Table 1).

**Table 1.** Baseline characteristics of NAKO/IMMEDIATE participants from Berlin-North study center.

| Baseline variables | N (%) or Mean (SD) or Median (IQR 25<sup>th</sup>,75<sup>th</sup> percentiles) |
| --- | --- |
| Age, years | 50.0 (43.0, 59.0) |
| Females | 1,288 (48.6) |
| Males | 1,362 (51.4) |
| Age category |  |
| 20-29 | 72 (2.7) |
| 30-39 | 260 (9.8) |
| 40-49 | 409 (15.4) |
| 50-59 | 922 (34.8) |
| 60-69 | 683 (25.8) |
| ≥ 70 | 304 (11.5) |
| Educational attainment* |  |
| ISCED 97 Low | 14 (0.6) |
| ISCED 97 Medium | 796 (32.5) |
| ISCED 97 High | 1,587 (64.8) |
| Enrolled in education | 52 (2.1) |
| Alcohol consumption* |  |
| Never | 66 (2.5) |
| Former | 78 (3.0) |
| Current 0 - < 5 g/day | 1,203 (45.9) |
| Current 5 - < 15 g/day | 701 (26.7) |
| Current ≥ 15 g/day | 574 (21.9) |
| Smoking status* |  |
| Never | 1,324 (50.5) |
| Former | 857 (32.7) |
| Current | 443 (16.9) |
| Body Mass Index, kg/m<sup>2</sup> | 25.5 (23.0,28.5) |
| Waist Circumference<sup>†</sup>, cm | 88.8 (80.1,98.3) |
| Hypertension<sup>†,‡</sup> | 739 (28.0) |
| Diabetes Mellitus<sup>†,‡</sup> | 137 (5.2) |
| Kidney Disease<sup>†,‡</sup> | 38 (1.4) |

\* ≤10.0% are missing
<sup>†</sup> ≤1.1% are missing
<sup>‡</sup> self-reported diagnosis
ISCED 97: International Standard Classification of Education 1997

Mean baseline values of HbA1c, PWV, SBP and eGFR were 34.6±5.8 mmol/mol, 8.30±1.53 m/s, 128.0±15.7 mmHg and 96.0±14.6 mL/min/1.73 m^2^, respectively (Table 2). CKM indicator mean values by sex are described in Table S2.

**Table 2.**
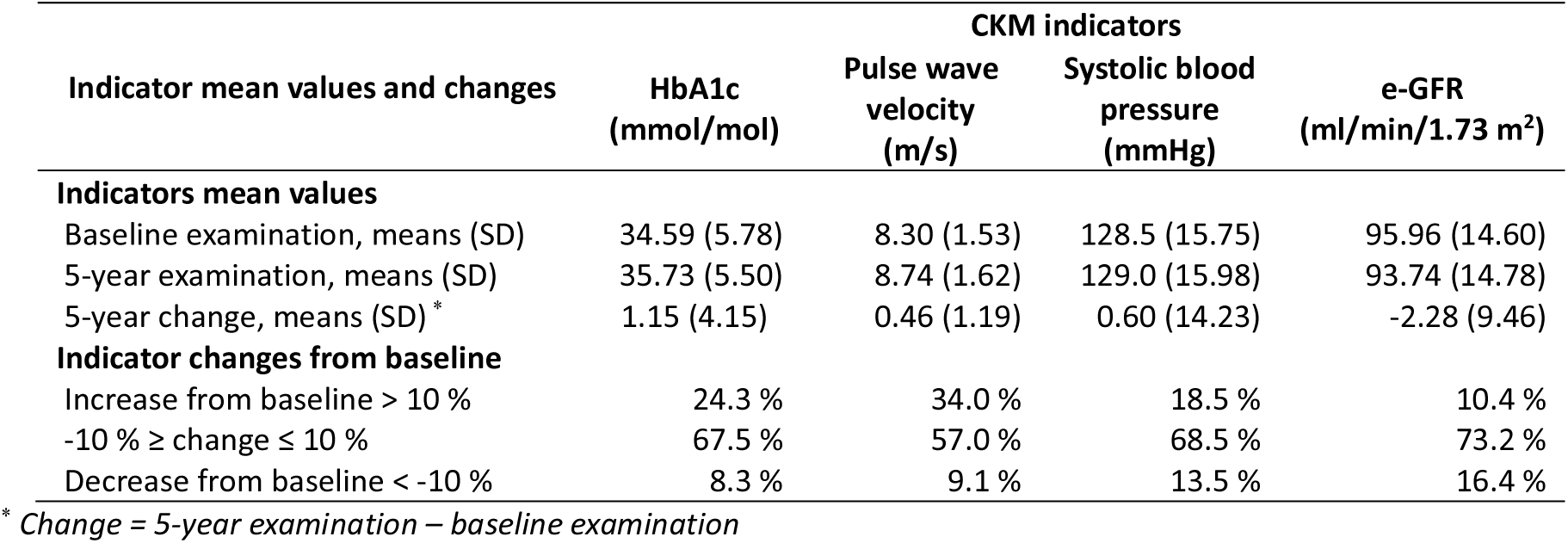
Baseline examination, 5-year examination, 5-year mean changes with standard deviations (SD) and number of participants (%) by categories of changes in HbA1c, pulse wave velocity, systolic blood pressure and e-GFR.

| Indicator mean values and changes | CKM indicators |  |  |  |
| --- | --- | --- | --- | --- |
|  | HbA1c<br>(mmol/mol) | Pulse wave<br>velocity<br>(m/s) | Systolic blood<br>pressure<br>(mmHg) | e-GFR<br>(ml/min/1.73 m<sup>2</sup>) |
| **Indicators mean values** |  |  |  |  |
| Baseline examination, means (SD) | 34.59 (5.78) | 8.30 (1.53) | 128.5 (15.75) | 95.96 (14.60) |
| 5-year examination, means (SD) | 35.73 (5.50) | 8.74 (1.62) | 129.0 (15.98) | 93.74 (14.78) |
| 5-year change, means (SD) * | 1.15 (4.15) | 0.46 (1.19) | 0.60 (14.23) | -2.28 (9.46) |
| **Indicator changes from baseline** |  |  |  |  |
| Increase from baseline > 10 % | 24.3 % | 34.0 % | 18.5 % | 10.4 % |
| -10 % ≥ change ≤ 10 % | 67.5 % | 57.0 % | 68.5 % | 73.2 % |
| Decrease from baseline < -10 % | 8.3 % | 9.1 % | 13.5 % | 16.4 % |

\* Change = 5-year examination – baseline examination

### Cross-sectional models in main analyses

In mutually adjusted cross-sectional analyses, higher age and higher BMI were each associated with higher HbA1c, PWV, and SBP, and with lower eGFR (Table 3, Table S3). For example, each 5-year higher baseline age was related to a 0.81 mmol/mol higher HbA1c (95% CI, 0.71,0.91), 0.36 m/s higher PWV (95% CI, 0.34,0.38), 1.79 mmHg higher SBP (95% CI, 1.51,2.06), and −4.13 mL/min/1.73m^2^ lower eGFR (95% CI, −4.34, −3.92). Higher WC residuals (i.e., WC adjusted for BMI) were associated with higher HbA1c, PWV, and SBP. Females versus males had lower PWV and SBP.

**Table 3.** Cross-sectional associations between the selected classical risk factors and the four CKM indicators, non-adjusted 95% confidence intervals and type 3 effect adjusted P values for categorical variables.

| Risk Factor | Difference in CKM indicators per difference in risk factor variables* |  |  |  |  |  |  |  |
| --- | --- | --- | --- | --- | --- | --- | --- | --- |
|  | HbA1c | Padj | PWV | Padj | SBP | Padj | eGFR | Padj |
|  | N = 2,642 |  | N = 2,647 |  | N = 2,647 |  | N = 2,633 |  |
| Age, per 5-years | 0.81 (0.71,0.91) | **<0.01** | 0.36 (0.34,0.38) | **<0.01** | 1.79 (1.51,2.06) | **<0.01** | -4.13 (-4.34,-3.92) | **<0.01** |
| Sex |  | 0.29 |  | **0.03** |  | **<0.01** |  | 0.88 |
| Male | Ref. |  | Ref. |  | Ref. |  | Ref. |  |
| Female | 0.31 (-0.27,0.90) |  | -0.13 (-0.25,-0.01) |  | -4.74 (-6.27,-3.21) |  | -0.11 (-1.25,1.04) |  |
| Educational attainment |  | **0.05** |  | **0.01** |  | 0.08 |  | 0.64 |
| ISCED 97 Low | 0.53 (-2.82,3.33) |  | 0.83 (0.09,1.56) |  | 3.21 (-6.79,13.2) |  | 2.98 (-1.84,7.78) |  |
| ISCED 97 Medium | 0.10 (-0.36,0.57) |  | 0.03 (-0.06,0.13) |  | 1.23 (0.03,2.43) |  | 0.19 (-0.73,1.10) |  |
| ISCED 97 High | Ref. |  | Ref. |  | Ref. |  | Ref. |  |
| Enrolled in education | 1.56 (0.51,2.62) |  | 0.27 (0.05,0.49) |  | 3.18 (0.02, 6.34) |  | 0.26 (-3.06,3.57) |  |
| Alcohol consumption |  | **0.02** |  | 0.11 |  | 0.33 |  | 0.45 |
| Never | 0.18 (-0.81,1.18) |  | 0.22 (0.02,0.43) |  | -0.56 (-3.81,2.69) |  | 0.50 (-2.42,3.41) |  |
| Former | 0.33 (-0.79,1.45) |  | 0.16 (-0.11,0.43) |  | -1.45 (-4.32,1.43) |  | 0.85 (-1.71,3.40) |  |
| Current 0 - < 5 g/day | Ref. |  | Ref. |  | Ref. |  | Ref. |  |
| Current 5 - < 15 g/day | -0.69 (-1.19,-0.20) |  | 0.05 (-0.06,0.16) |  | 0.94 (-0.40,2.28) |  | 0.81 (-0.25,1.87) |  |
| Current ≥ 15 g/day | -0.62 (-1.18,-0.06) |  | 0.08 (-0.05,0.20) |  | 0.40 (-1.10,1.91) |  | 1.19 (0.10,2.28) |  |
| Smoking status |  | **<0.01** |  | **<0.01** |  | **<0.01** |  | **0.02** |
| Never | Ref. |  | Ref. |  | Ref. |  | Ref. |  |
| Former | -0.02 (-0.50,0.45) |  | -0.08 (-0.19,0.02) |  | -2.16 (-3.43,-0.89) |  | -0.40 (-1.37,0.57) |  |
| Current | 1.47 (0.90,2.04) |  | -0.20 (-0.32,-0.09) |  | -3.44 (-4.94,-1.94) |  | 1.60 (0.44,2.75) |  |
| Body Mass Index, kg/m<sup>2</sup> | 0.24 (0.18,0.30) | **<0.01** | 0.03 (0.02,0.04) | **<0.01** | 0.75 (0.62,0.89) | **<0.01** | -0.14 (-0.24,-0.04) | **0.04** |
| WC residuals, cm | 0.10 (0.05,0.15) | **<0.01** | 0.03 (0.02,0.04) | **<0.01** | 0.15 (0.03,0.27) | **0.02** | 0.10 (0.01,0.19) | 0.09 |

Results obtained from separate multivariable linear regression models for each outcome variable.
Multivariable regression models include all predictor variables. Significant associations are marked in bold. Confidence intervals are not adjusted for multiple testing.
\* Results can be interpreted as the difference in the CKM indicators that is observed for a specified difference in the predictor. For example, 5-year higher age is related to a 0.81 mmol/mol (95%-CI 0.71,0.91) higher HbA1c concentration, while women compared to men have 0.13 m/s lower PWV (-0.25,-0.01).
P values are adjusted using the Benjamini-Hochberg procedure.

Lower education was associated with higher HbA1c and with higher PWV. Higher alcohol consumption was related to lower HbA1c. Current smoking was associated with higher HbA1c and eGFR and with lower PWV and SBP.

### Cross-sectional models in sensitivity analyses

In sex-stratified analyses with tests for interaction by sex, none of the associations were significantly different between males and females (all p-interaction ≥0.09), with the exception of the association of age with SBP, which was significantly stronger in females (2.60; 95%-CI 2.24-2.95 mmHg per 5 years) compared to men (0.95; 95%-CI 0.54-1.36 mmHg per 5 years, p-interaction <0.01) (Tables S4).

In models controlled for diabetes and hypertension medication, associations between sex and PWV, between educational attainment and HbA1c or PWV, between alcohol consumption and HbA1c, between smoking or BMI and eGFR were attenuated (Table S5). In models controlled for physical activity per day the association between sex and PWV was attenuated (Table S6).

### Longitudinal models in main analyses

Mean follow-up time between baseline and second examination was 4.92±0.48 years. Over 5 years, HbA1c, PWV and SBP increased on average by 1.15±4.15 mmol/mol (+3.3% from baseline), 0.46±1.19 (+5.3% from baseline) and 0.60±14.23 mmHg (+0.8% from baseline), respectively, whereas eGFR decreased on average by −2.28±9.46 mL/min/1.73 m^2^ (−2.4% from baseline). Over 5 years, CKM indicators changed less than 10% from baseline values for most participants. The PWV and SBP were the indicators with the highest proportion of decline and highest change variability, respectively (Table2). CKM indicator mean absolute and relative changes by sex are described in Table S2.

Age and BMI were associated with the magnitude of change in PWV over 5 years (β = 0.03 m/s per 5 year of age, 95% CI 0.01, 0.06 m/s and β = −0.02 m/s per 1kg/m^2^, 95% CI −0.03, −0.01 m/s, respectively), with older age associated with greater increases in PWV and higher BMI associated with lower increases in PWV. None of the other risk factors under investigation were associated with prospective changes in the studied CKM indicators (Tables 4, S7).

**Table 4.** Longitudinal associations between the selected classical risk factors and the four CKM indicator changes, non-adjusted 95% confidence intervals and type 3 effect adjusted P values for categorical variables.

| Risk Factor | 5-year change in CKM function per difference in factor variables* (95% CI) |  |  |  |  |  |  |  |
| --- | --- | --- | --- | --- | --- | --- | --- | --- |
|  | HbA1c | Padj | PWV | Padj | SBP | Padj | eGFR | Padj |
|  | N = 2,642 |  | N = 2,647 |  | N = 2,647 |  | N = 2,633 |  |
| Age, per 5-years | 0.01 (-0.06,0.09) | 0.97 | **0.03 (0.01,0.06)** | **0.04** | -0.16 (-0.43,0.11) | 0.52 | -0.04 (-0.23,0.14) | 0.93 |
| Sex |  |  |  |  |  |  |  |  |
| Male | Ref. | 0.98 | Ref. | 0.22 | Ref. | 0.37 | Ref. | 0.77 |
| Female | 0.00 (-0.45,0.46) |  | -0.09 (-0.21,0.03) |  | 0.76 (-0.74,2.27) |  | -0.16 (-1.20,0.88) |  |
| Educational attainment |  |  |  |  |  |  |  |  |
| ISCED 97 Low | 0.68 (-0.44,1.81) | 0.98 | -1.14 (-2.13,-0.15) | 0.18 | -7.89 (-14.8,-0.94) | 0.25 | -2.32 (-6.13,1.49) | 0.77 |
| ISCED 97 Medium | 0.11 (-0.25,0.47) |  | 0.02 (-0.08,0.12) |  | -0.11 (-1.32,1.11) |  | -0.11 (-0.92,0.71) |  |
| ISCED 97 High | Ref. |  | Ref. |  | Ref. |  | Ref. |  |
| Enrolled in education | -0.07 (-0.93,0.80) |  | 0.16 (-0.11,0.43) |  | 0.23 (-3.04, 3.50) |  | 0.38 (-2.74,3.51) |  |
| Alcohol consumption |  |  |  |  |  |  |  |  |
| Never | 0.08 (-0.83,1.00) | 0.98 | 0.10 (-0.14,0.34) | 0.20 | 0.05 (-3.07,3.17) | 0.25 | -1.73 (-4.10,0.64) | 0.77 |
| Former | 0.72 (-0.24,1.67) |  | -0.17 (-0.41,0.07) |  | 1.49 (-1.41,4.39) |  | -0.68 (-2.74,1.39) |  |
| Current 0 - < 5 g/day | Ref. |  | Ref. |  | Ref. |  | Ref. |  |
| Current 5 - < 15 g/day | 0.04 (-0.36,0.45) |  | 0.06 (-0.05,0.17) |  | 0.34 (-0.98,1.66) |  | -0.11 (-1.04,0.81) |  |
| Current ≥ 15 g/day | -0.17 (-0.63,0.28) |  | 0.12 (-0.00,0.25) |  | 2.00 (0.43,3.57) |  | 0.54 (-0.43,1.51) |  |
| Smoking status |  |  |  |  |  |  |  |  |
| Never | Ref. | 0.98 | Ref. | 0.54 | Ref. | 0.27 | Ref. | 0.77 |
| Former | 0.09 (-0.29,0.46) |  | 0.07 (-0.04,0.18) |  | 1.02 (-0.26,2.29) |  | -0.01 (-0.84,0.82) |  |
| Current | 0.06 (-0.40,0.52) |  | 0.03 (-0.09,0.14) |  | 1.18 (-0.37,2.73) |  | -0.85 (-1.97,0.25) |  |
| Body Mass Index, kg/m<sup>2</sup> | 0.05 (-0.00,0.10) | 0.68 | **-0.02 (-0.03,-0.01)** | **<0.01** | -0.12 (-0.26,0.02) | 0.35 | 0.01 (-0.07,0.10) | 0.93 |
| WC residuals, cm | 0.00 (-0.04,0.04) | 0.97 | -0.00 (-0.01,0.01) | 0.73 | 0.00 (-0.12,0.12) | 0.97 | -0.04 (-0.12,0.04) | 0.84 |

Results obtained from separate multivariable linear regression models for each change in outcome variables.
Multivariable regression models include all predictor variables. Outcome variables were calculated as the participants follow-up value minus baseline value, divided by the length of participants' follow-up, multiplied by 5 years. Nominal significant associations are marked in bold with non-adjusted confidence intervals, as well as adjusted p-values (Padj) <0.05.
\* Results can be interpreted as the change in the CKM indicators over 5 years beyond the average change that is observed for a specified difference in the predictor. For example, 5-year higher age at baseline is related to a 0.03 m/s (95%-CI 0.01-0.06) stronger increase in PWV. P values are adjusted using the Benjamini-Hochberg procedure.

### Longitudinal models in sensitivity analyses

In sex-stratified analyses with tests for interaction by sex, none of the associations were significantly different between males and females (Tables S8).

In models controlled for diabetes and hypertension medication, the association between age and PWV 5-year changes was attenuated (Table S9). In models adjusted for daily physical activity, the associations of age and BMI with 5-year changes in PWV remained similar in magnitude and direction to those observed in the main analysis, although the corresponding adjusted P values were attenuated and no longer below 0.05 (Table S10).

## Discussion

### Main findings

In this study, different associations were observed between baseline classical risk factors and baseline CKM indicators (cross-sectional analyses) and between baseline classical risk factors and CKM indicator 5-year changes (longitudinal analyses). Cross-sectionally, participants with higher age and higher BMI had higher levels of SBP, HbA1c, PWV and lower levels of eGFR. Moreover, female versus male participants had lower PWV and SBP. Alcohol consumption was associated with higher PWV and with lower HbA1c levels. Smoking was associated with higher HbA1c and eGFR levels and with lower PWV and SBP levels. Higher WC residuals were associated with higher HbA1c, PWV, and SBP. Longitudinally, SBP, HbA1c and PWV slightly increased over 5-years, and eGFR decreased. While higher age was associated with a stronger 5-year increase in PWV and higher BMI was associated with a lower PWV increase, none of the other classical risk factors at baseline was significantly related to 5-year changes in CKM indicators. These data suggest that although most of these classical risk factors are related to CKM indicators, only very few predict 5-year changes.

### Comparisons with other studies

As described in other cross-sectional studies, age and BMI were consistently associated with the 4 CKM indicators, namely with higher levels of HbA1c, PWV, SBP or with lower eGFR levels, confirming their major role in determining CKM function^1, 2^. Other classical risk factors were less consistently associated with all baseline CKM indicators, despite CKM diseases being highly intertwined. For instance, as reported in the literature, females compared to males had lower SBP^5^ and lower PWV^4^, but neither lower HbA1c nor higher eGFR. In the same line, while smoking was positively associated with HbA1c, as previously reported^7^, it was negatively associated with SBP and PWV and positively associated with eGFR. These heterogeneous associations may relate to observed changing sex/HbA1c associations across age groups^35^ or to the lower BMI, and hence lower BP, reported in smokers *vs* non smokers^36^. Finally, lack of association between sex and eGFR or a positive association between smoking and eGFR may relate to the more frequent proteinuria, and hence glomerular hyperfiltration rate, in males *vs* females and in smokers *vs* non smokers^14, 37^. Therefore, in cross-sectional analyses, CKM indicators share many classical risk factors although probable confounders may attenuate or reverse expected associations.

Over the 5 years of follow-up, baseline HbA1c, PWV and SBP mean levels increased by 3.3%, 5.3% and 0.8% and baseline eGFR mean levels decreased by 2.4%. These mild changes were in line with the expected slow aging process of a middle-aged general population^3, 4, 8, 38^. For most participants, CKM indicator changed less than 10%, an approximate threshold often used at the individual level to differentiate biologically significant changes between two consecutive measurements from other sources of variability (e.g., within-subject biological coefficient of variation and analytical coefficient of variation)^30–32^. However, although most participants may have had only mild sub clinical changes, the changes on a population level are still relevant to consider in studying health-to-disease transitions. The relative change in CKM indicators was strongest for the increase in PWV, in line with being an accurate indicator of ageing^28^ and lowest for the increase in SBP. Notably, although CKM indicators at the population average deteriorated, at the individual level, several persons also had improvements in CKM indicators. This may reflect true improvements in organ function, regression to the mean^33^, or, in the case of eGFR values, increased glomerular hyperfiltration due to creatininuria in participants transitioning towards chronic kidney disease^38^.

Although classical risk factors were shared across CKM indicators cross-sectionally, in our analysis, only age and BMI were significantly related to changes in PWV, whereas no other associations of classical risk factors with longitudinal changes were observed. While higher age was related to stronger 5-year increases in PWV, higher BMI was related to lower increases. This suggests that – except for the relationship of age and BMI to changes in PWV – participants with any given risk factor have higher or lower CKM indicators but these risk factors do not predict changes in these indicators beyond the average change that is observed in the population. For example, persons with higher BMI have higher HbA1c concentrations compared to persons with lower BMI, however, the 5-year increase in HbA1c is not different between these persons.

While higher age was related to stronger 5-year increases in PWV, higher BMI was related to lower increases. These data suggest that older compared to younger people not only have higher PWV but also a stronger PWV increase over 5-years. Previous studies found similar results^4, 36, 37^. Of note, this association was attenuated after adjustment for antihypertensive and antidiabetic medication, suggesting that the association between age and vascular stiffness progression may be partly influenced by treatment-related factors. Persons with higher BMI have higher PWV compared to those with lower BMI, but, counter-intuitively, they have a lower 5-year increase, as described in other studies^39, 40^. It has been previously reported that, although mean effect of BMI on certain cardiovascular outcomes increases with age (cross-sectional effect), this effect decelerates with age (longitudinal effect)^41^. This may relate to a stronger detrimental effect of BMI on certain cardiovascular outcomes in younger ages; while in older ages, higher BMI may be an indicator of a higher lean mass and less sarcopenia, and therefore a less detrimental effect of BMI on the cardiovascular outcomes^42^. Notably, physical activity is known to attenuate lean mass decline with age^43^, which may partly contribute to the attenuated BMI/PWV changes association in models controlled for physical activity.

### Strengths and limitations

Strengths of the present study are the combination of the cross-sectional and the longitudinal design and the use of highly standardized and quality-controlled variables within the NAKO-cohort, which allows a comprehensive comparison of associations across study designs. A limitation is that our change-model is currently only based on 2 time points, thus limiting the change model to a linear design, and restricted to a 5-year period. A second follow up examination after 10 years within the NAKO is currently ongoing, which will allow to model non-linear changes over longer time-periods. The IMMEDIATE sub-sample is a phenotypically enriched and selected subsample of limited size probably limiting the generalizability of the results. However, participants do not differ in their baseline socio-demographic, phenotypic and lifestyle characteristics from the general NAKO cohort^44^. At a methodological level, different results found between study designs may result from cohort effect described in cross-sectional designs and/or to the lower statistical power described in longitudinal designs^45^.

## Conclusion

In conclusion, we found in our analysis most of the classical risk factors related to many of the CKM indicators cross-sectionally, but only age and BMI related to changes in PWV, whereas no other associations of classical risk factors with longitudinal changes were observed. These data suggest that classical CKM risk factors such as age, sex, educational attainment, alcohol and tobacco consumption, BMI and WC have greater utility for assessing cross-sectional CKM function than for predicting prospective changes in CKM function in the general population.

## Data Availability

Due to privacy concerns, the data supporting this study cannot be shared publicly but can be accessed with approval from the institutional review board and upon reasonable request

## Acknowledgments

We thank all participants of the German National Cohort (NAKO) and the staff of this research initiative. Members and affiliations of the NAKO Investigator Consortium can be accessed via www.nako.de/principal-investigators.

## Non-standard Abbreviations and Acronyms

baPTT: brachial-ankle pulse transit time
BMI: Body Mass Index
CKD-EPI: Chronic Kidney Disease Epidemiology Collaboration
CKM: cardio-Kidney and metabolic
eGFR: estimated Glomerular Filtration rate
HbA1c: Glycated hemoglobin A1c
IMMEDIATE: Imminent Disease Prediction and Prevention at the Environment Host Interface
ISCED 97: International Standard Classification of Education 1997
NAKO: German national Cohort
PWV: Pulse Wave Velocity
SBP: Systolic Blood pressure
WC: Waist Circumference

---

**Table S1** Baseline characteristics of NAKO/IMMEDIATE participants from study center Berlin-North by sex

**Table S2** Baseline examination, 5-year examination, 5-year mean changes with standard deviations (SD) and number of participants (%) by categories of changes in HbA1c, pulse wave velocity, systolic blood pressure and e-GFR

**Table S3** Cross-sectional associations between classical factors and four CKM indicators, non-adjusted 95% confidence intervals and adjusted P values with the Benjamini-Hochberg procedure

**Table S4** Cross-sectional associations between classical risk factors and the four CKM indicators stratified by sex and with adjusted values of sex*risk factor interaction terms

**Table S5** Cross-sectional associations between classical risk factors and the four CKM indicators controlling for diabetes and hypertension medication. P values are adjusted using the Benjamini-Hochberg procedure

**Table S6** Cross-sectional associations between classical risk factors and the four CKM indicators controlling for physical activity per day. P values are adjusted using the Benjamini-Hochberg procedure

**Table S7** Longitudinal associations between risk factors and four CKM indicators, non-adjusted 95% confidence intervals and adjusted P values using the Benjamini-Hochberg procedure

**Table S8** Longitudinal associations between classical risk factors and the four CKM indicators stratified by sex and with adjusted values of sex*risk factor interaction terms

**Table S9** Longitudinal associations between classical risk factors and the four CKM indicators controlling for diabetes and hypertension medication. P values are adjusted using the Benjamini-Hochberg procedure

**Table S10** Longitudinal associations between classical risk factors and the four CKM indicators controlling for physical activity per day. P values are adjusted using the Benjamini-Hochberg procedure

